# Association study of HLA with the kinetics of SARS-CoV-2 spike specific IgG antibody responses to BNT162b2 mRNA vaccine

**DOI:** 10.1101/2022.02.01.22270285

**Authors:** Seik-Soon Khor, Yosuke Omae, Junko S. Takeuchi, Ami Fukunaga, Shohei Yamamoto, Akihito Tanaka, Kouki Matsuda, Moto Kimura, Kenji Maeda, Gohzoh Ueda, Tetsuya Mizoue, Mugen Ujiie, Hiroaki Mitsuya, Norio Ohmagari, Wataru Sugiura, Katsushi Tokunaga

## Abstract

BNT162b2, an mRNA-based SARS-CoV-2 vaccine (Pfizer-BioNTech), is one of the most effective COVID-19 vaccines and has been approved by more than 130 countries worldwide. However, several studies have reported that the COVID-19 vaccine shows high interpersonal variability in terms of humoral and cellular responses, such as those with respect to SARS-CoV-2 spike protein immunoglobulin (Ig)G, IgA, IgM, neutralizing antibodies, and CD4^+^ & CD8^+^ T cells. The objective of this study is to investigate the kinetic changes in anti-SARS-CoV-2 spike IgG (IgG-S) profiles and adverse reactions and their associations with HLA profiles among 100 hospital workers from the Center Hospital of the National Center for Global Health and Medicine (NCGM), Tokyo, Japan. DQA1*03:03:01 (P = 0.017; Odd ratio (OR) 2.80, 95%Confidence interval (CI) 1.05–7.25) was significantly associated with higher IgG-S production after two doses of BNT162b2 while DQB1*06:01:01:01 (P = 0.028, OR 0.27, 95%CI 0.05–0.94) was significantly associated with IgG-S declines after two doses of BNT162b2. No HLA alleles were significantly associated with either local symptoms or fever. However, C*12:02:02 (P = 0.058; OR 0.42, 95%CI 0.15–1.16), B*52:01:01 (P = 0.031; OR 0.38, 95%CI 0.14–1.03), DQA1*03:02:01 (P = 0.028; OR 0.39, 95%CI 0.15–1.00) and DPB1*02:01:02 (P = 0.024; OR 0.45, 95%CI 0.21–0.97) appeared significantly associated with protection against systemic symptoms after two doses of BNT162b2 vaccination. Further studies with larger sample sizes are clearly warranted to determine HLA allele associations with the production and long-term sustainability of IgG-S after COVID-19 vaccination.

## Introduction

The concept of messenger RNA (mRNA) vaccination stems from 1987, when Robert Malone confirmed that human cells can absorb cationic liposomes containing mRNA and can create proteins from those mRNA sequences (1). However, over the years, academic laboratories and companies working on mRNA had come to the consensus that mRNA is too prone to degradation to be used effectively as a drug or a vaccine(2). From the end of the 20^th^ century, research into mRNA vaccines mainly focused on influenza disease (3) and cancer (4), with testing in animal models yielding satisfactory results. Such results inspired CureVac, BioNTech, and Moderna to focus on transforming mRNA into a drug platform (5-7). In 2005, the Karikó and Weissman team performed a landmark experiment in which an mRNA vaccine was used to successfully suppress RNA recognition by Toll-like receptors (TLRs) using modified pseudouridine, a uridine analog (8). All this previous work contributed to the rapid development of coronavirus disease 2019 (COVID-19) mRNA vaccines within days of the genome for the severe acute respiratory syndrome coronavirus 2 (SARS-CoV-2) being released online(9).

BNT162b2, an mRNA-based SARS-CoV-2 vaccine (Pfizer-BioNTech), is one of the most effective COVID-19 vaccines (10) and has been approved by more than 130 countries worldwide. However, several studies have reported that the COVID-19 vaccine shows high interpersonal variability in terms of humoral and cellular responses, such as those with respect to anti-SARS-CoV-2 spike immunoglobulin (Ig)G (11-17), IgA (15, 16), IgM (16), neutralizing antibodies (18-20), and CD4^+^ & CD8^+^ T cells (18, 19, 21). The long-term sustainability of SARS-CoV-2 neutralizing antibodies is crucial for the protection against infection and hospitalization.

COVID-19 mRNA vaccines work by introducing the SARS-CoV-2 spike protein (S-protein) mRNA sequence and subsequently the translated protein to the human immune system. This first prompts an innate (non-specific) immune response by triggering immune cells such as natural killer cells, macrophages, neutrophils, dendritic cells, mast cells, basophils, and eosinophils. The second line of immune response is the adaptive immune response, in which T and B lymphocytes are activated to generate long-term specific immune responses to the SARS-CoV-2 virus.

Human Leukocyte Antigen (HLA) allelic variation may be associated with vaccine efficacy according to trials of hepatitis B vaccine (22, 23), influenza vaccine (24) and HIV-1 vaccine (25). However, data on possible associations with the kinetics of post-COVID-19 vaccination antibodies remain scarce. Here, we designed a longitudinal study aimed at investigating the association of host HLA polymorphisms with kinetic changes in IgG and adverse reactions during and after BNT162b2 vaccination among hospital workers at a national medical institution in Japan.

## Methods

### Study design and participants

The National Center for Global Health and Medicine (NCGM) is a Japanese government-designated medical center for the treatment of COVID-19 patients. We recruited and monitored a total of 100 hospital workers ≥ 20 years of age from the period between March and June 2021. Details of the study design are explained in previous studies(26, 27). All participants were vaccinated with two doses of the BNT162b2 mRNA-based SARS-CoV-2 vaccine (Pfizer-BioNTech) according to the standard protocol (two doses of 30 µg, administered 3 weeks apart) (18, 28). All participants received the first dose of the BNT162b2 vaccine in March 2021 and the second dose was administered 21 days after the first dose. Blood was drawn from volunteers on day 1 (immediately after the first dose). Samples were then collected on day 15, day 29 (7 days after the second dose) and day 61. None of the participants had any history of COVID-19 or administration of immunosuppressive medications. The study protocol was approved by the Ethics Committee of the NCGM, Japan (approval number: NCGM-A-004175-00). Written informed consent was obtained from all participants prior to enrollment.

### Self-reported questionnaire

Participants were requested to complete a questionnaire regarding the adverse reactions experienced after each dose of vaccine. Based on the results, we classified side effects into local symptoms (pain at the injection site, swelling and redness) and systemic symptoms (fever, dizziness, fatigue, headache, chills, vomiting, diarrhea, muscle pain and joint pain) with reference to the U.S. Food and Drug Administration guidance. Symptoms were further graded into 4 categories: grade 0, no adverse effect; grade 1, adverse effect without interfering with daily activities; grade 2, adverse effect with some degree of interference with daily activities; and grade 3, adverse effect with considerable interference with daily activities. Redness or swelling at the side of infection was graded based on the size of the redness or swelling as: grade 0, 0–2.0 cm; grade 1, 2.1–5.0 cm; grade 2, 5.1–10.0 cm; and grade 3, >10.0 cm.

### Serological assays

Antibodies against the SARS-CoV-2 receptor-binding domain (RBD) of the S1 subunit of the spike protein (IgG-S) (AdviseDx SARS-CoV-2 IgG II; Abbott, Chicago, IL) and SARS-CoV-2 nucleocapsid (IgG-N) (Abbott ARCHITECT® SARS-CoV-2 anti-N IgG; Abbott) were measured at each collection time point. For IgG-S, results > 50 AU/mL (as the cut-off set by the manufacturer) were considered indicative of seropositivity. For surrogate measurement of neutralizing antibody, a threshold of 4160 AU/mL was applied, as this threshold corresponds to a 95% probability of obtaining a positive result from plaque reduction neutralization test (PRNT) for SARS-CoV-2 at 1:250 dilution (14). For IgG-N, results above the index value of 1.40 (as the cut-off set by the manufacturer) were considered indicative of seropositivity.

#### Next-generation sequencing (NGS)-based HLA genotyping

NGS HLA genotyping was performed using AllType™ NGS Assays (One Lambda, West Hills, CA) on an Ion GeneStudio S5 sequencing system (Thermo Fisher Scientific, Waltham, MA). HLA-A, -C, -B, DRB1, DQA1, DQB1, DPA1, DPB1 targeted gene amplification, HLA library preparation, HLA template preparation, and HLA library loading onto an ion 530v1 chip in an Ion Chef library preparation robot (Thermo Fisher Scientific) and final sequencing in an Ion GeneStudio S5 sequencer (Thermo Fisher Scientific) were performed following the instructions from the vendor.

#### HLA genotype assignment

Demultiplexing of barcodes and base-calling was carried out using Torrent Suite version 5.8.0 (Thermo Fisher Scientific). Raw fastq reads were extracted using the FileExporter function in Torrent Suite version 5.8.0. HLA genotype assignments were undertaken using HLATypeStream Visual (TSV v2.0; One Lambda, West Hills, CA) and NGSengine® (v2.22.0.22581; GenDX, Utrecht, Netherlands). The default analysis parameters and healthy metrics threshold were applied for TSV v2.0, while we applied the “ignore regions” function in NGSengine® to eliminate known sequencing error sites in the ion S5 system. Four-field resolution HLA alleles were obtained for HLA-A, -C, -B, DRB1, DQA1, DQB1, DPA1. Novel HLA alleles were subjected to Pacbio Sequel sequencing in collaboration with the H.U. Group Research Institute (Tokyo, Japan). Pacbio subreads and consensus reads were obtained from SMRT Link software and HLA calling was performed using NGSengine® software (v2.22.0.22581; GenDX).

### Statistical analysis

Correlation coefficient tests and Kruskal-Wallis tests were applied to describe the correlation between IgG-S titers with age, sex, BMI of participants, and adverse reactions after two doses of vaccination. IgG-S levels in participants plateaued on day 29 (7 days after the second dose), and were further classified into low responders (top 25^th^ percentile of the IgG-S distribution) and high responders (bottom <25^th^ percentile of the IgG-S distribution). Strong decliners (top 25^th^ percentile of Δ) and weak decliners (bottom 75^th^ percentile of Δ) were defined as Δ of IgG-S level between day 29 (7 days after the second dose) and day 61 (39 days after second vaccination).

Case-control HLA allele association tests, HLA haplotype estimations and case-control HLA haplotype association tests were prepared and analyzed using the Bridging ImmunoGenomics Data Analysis Workflow Gaps (BIGDAWG) R package (29).

## Results

### Basic characteristics of participants

A total of 100 hospital workers at the NCGM were recruited, comprising 32 men and 68 women. Participants were further classified into 5 age strata to observe the changes of IgG-S in relation to age: 20–29 years old; 30–39 years old; 40–49 years old; 50–59 years old; and <u>≥</u> 60 years old (Table 1). The BMI of participants ranged from 17 kg/m^2^ to 38 kg/m^2^, with the highest BMI observed in the group ≥ 60-years old (mean ±standard deviation, 26 ±6.30 kg/m^2^).

**Table 1.**
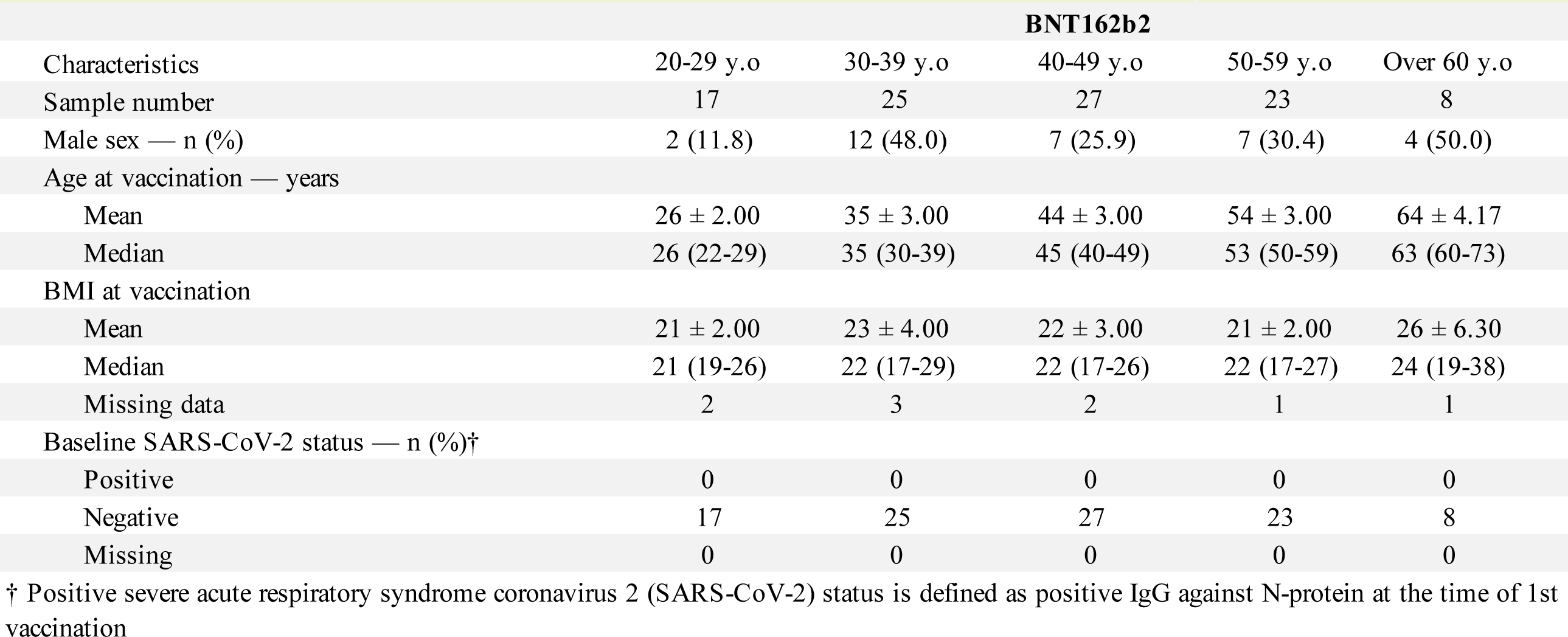
Basic characteristics of study participants.

| Table 1. Basic characteristics of study participants |  |  |  |  |  |
| --- | --- | --- | --- | --- | --- |
|  |  |  | BNT162b2 |  |  |
| Characteristics | 20-29 y.o | 30-39 y.o | 40-49 y.o | 50-59 y.o | Over 60 y.o |
| Sample number | 17 | 25 | 27 | 23 | 8 |
| Male sex — n (%) | 2 (11.8) | 12 (48.0) | 7 (25.9) | 7 (30.4) | 4 (50.0) |
| Age at vaccination — years |  |  |  |  |  |
| Mean | 26 ± 2.00 | 35 ± 3.00 | 44 ± 3.00 | 54 ± 3.00 | 64 ± 4.17 |
| Median | 26 (22-29) | 35 (30-39) | 45 (40-49) | 53 (50-59) | 63 (60-73) |
| BMI at vaccination |  |  |  |  |  |
| Mean | 21 ± 2.00 | 23 ± 4.00 | 22 ± 3.00 | 21 ± 2.00 | 26 ± 6.30 |
| Median | 21 (19-26) | 22 (17-29) | 22 (17-26) | 22 (17-27) | 24 (19-38) |
| Missing data | 2 | 3 | 2 | 1 | 1 |
| Baseline SARS-CoV-2 status — n (%)† |  |  |  |  |  |
| Positive | 0 | 0 | 0 | 0 | 0 |
| Negative | 17 | 25 | 27 | 23 | 8 |
| Missing | 0 | 0 | 0 | 0 | 0 |

† Positive severe acute respiratory syndrome coronavirus 2 (SARS-CoV-2) status is defined as positive IgG against N-protein at the time of 1st vaccination

### Kinetics of antibodies activities

#### IgG-S

All participants tested negative for IgG-S, IgG-N and IgM at day 1 (before BNT162b2 vaccination). IgG-S levels increased exponentially after the first dose of BNT162b2 vaccination, plateauing on day 15. After the second dose of BNT162b2 vaccination, the IgG-S level peaked on day 29 (7 days after the second vaccination). Younger age (especially under 59 years old) (Kruskal-Wallis test, P = 0.024) (Supplementary Figure 1a) and female sex (Kruskal-Wallis test, P = 0.034) (Supplementary Figure 1b) were significantly associated with higher IgG-S production after two doses of BNT162b2 vaccinations (Supplementary Figure 1a–c). For the surrogate measurement of neutralizing antibody activities, we examined for IgG-S level > 4160 AU/mL, which corresponded to a 95% probability of obtaining a PRNT ID50 (estimated number of virus particles required to produce infection in 50% of normal adult humans) at 1:250 dilution(14). Fifteen days after the first dose of BNT162b2 vaccination, none of the participants had reached a sufficient level of surrogate neutralizing activities, although 96% of participants obtained sufficient level of surrogate neutralizing activities against SARS-CoV-2 by day 29 (7 days after the second dose). However, the proportion showing sufficient level of surrogate neutralizing activities decreased to 81% on day 61.

At day 61 (39 days after second vaccination), we observed a significant correlation between age (r = 0.325, P = 0.0012) (Supplementary Figure 2a) and decreased IgG-S levels, but no associations with sex (P = 0.0621) (Supplementary Figure 2b) or BMI (r = 0.173, P = 0.1065) (Supplementary Figure 2c) were apparent for participants. However, although it is not statistically significant, strong decliners (<u>Δ</u> ≥20,000 AU/mL) were observed to be mainly concentrated among the female group (n=15, 23%), compared with the male group (n=3, 9.4%).

#### IgG-N

None of the participants were seropositive for IgG-N across the five survey time points, indicating no prior SARS-CoV-2 infection. No infection was observed during or after the BNT162b2 vaccination.

### Associations of HLA alleles with adverse reactions and IgG-S levels

We classified the self-reported body temperature into 4 categories: T1, < 37.5°C; T2, 37.5–37.9°C; T3, 38.0–38.4°C; and T4, 38.5–38.9°C. Systemic symptoms (grade 0–1 versus grade 2–3) showed a significant association (Kruskal-Wallis test, P = 0.026) with higher IgG-S level on day 29 (7 days after the second dose) (Supplementary Figure 3a). More specifically, fever (> 37.5°C) showed a strong association (Kruskal-Wallis test, P = 0.009) with higher IgG-S level on day 29 (Supplementary Figure 3b). However, local symptoms (Kruskal-Wallis test, P = 0.465) were not associated with IgG-S level on day 29 (Supplementary Figure 3c).

DQA1*03:03:01 (P = 0.017; OR 2.80, 95%CI 1.05–7.25) was significantly associated with higher IgG-S production after two doses of BNT162b2 (Table 2). Haplotype analysis of HLA-DQA1 and surrounding HLA genes did not reveal any significant associations with HLA haplotypes (DQA1*03:03:01-DQB1*04:01:01, P = 0.072, OR 2.45, 95%CI 0.92–6.52 and DRB1*04:05:01-DQA1*03:03:01-DQB1*04:01:01, P = 0.072, OR 2.45, 95%CI 0.92–6.52) (Table 3).

**Table 2.**
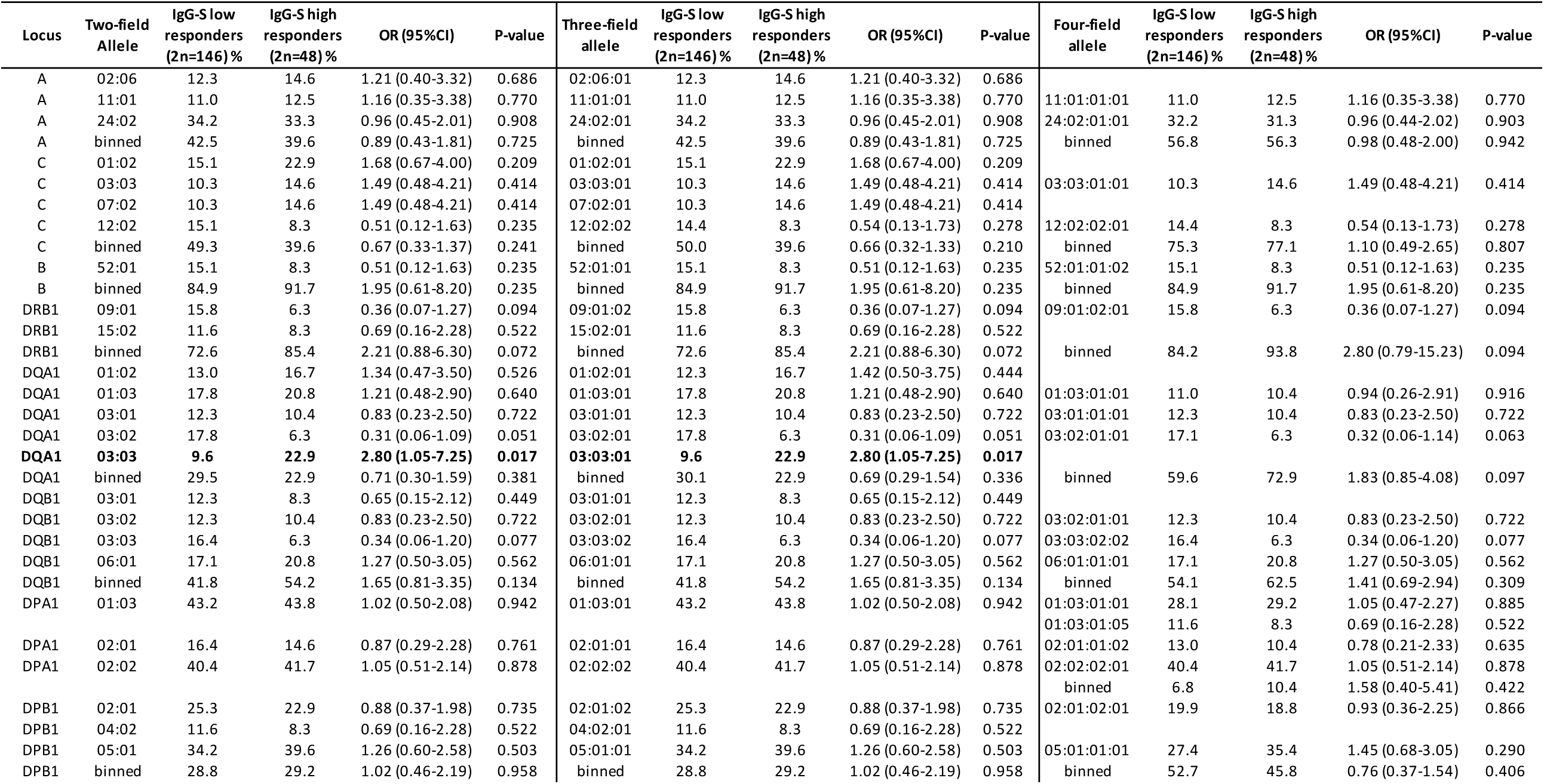
Associational analysis of HLA-A, -C, -B, -DRB1, -DQB1, -DOA1 and -DPB1 alleles in 73 IgG-S low responders and 24 IgG-S high responders. Significant HLA alleles are highlighted. Abbreviations: OR, odds ratio; 95%CI, 95% confidence interval; binned, rare HLA alleles with expected count < 5 are combined into a common class.

**Table 3.** HLA haplotype analysis for HLA-DRB1, -DQA1, and -DQB1 in IgG-S low responders versus high responders. Abbreviations: OR, odds ratio; 95%CI, 95% confidence Interval; binned, rare HLA alleles with expected count < 5 are combined into a common class.

| Locus | Allele | IgG-S low responders<br>(2n=146) % | IgG-S high responders<br>(2n=48) % | OR (95%CI) | P-value |
| --- | --- | --- | --- | --- | --- |
| DRB1~DQA1~DQB1 | 09:01:02~03:02:01~03:03:02 | 14.4 | 6.3 | 0.40 (0.07-1.43) | 0.138 |
| DRB1~DQA1~DQB1 | 15:02:01~01:03:01~06:01:01 | 11.0 | 8.3 | 0.74 (0.23-2.33) | 0.605 |
| DRB1~DQA1~DQB1 | 15:01:01~01:02:01~06:02:01 | 8.2 | 10.4 | 1.30 (0.43-3.89) | 0.641 |
| DRB1~DQA1~DQB1 | 04:05:01~03:03:01~04:01:01 | 7.5 | 16.7 | 2.45 (0.92-6.52) | 0.072 |
| DRB1~DQA1~DQB1 | 08:03:02~01:03:01~06:01:01 | 6.2 | 10.4 | 1.77 (0.56-5.57) | 0.329 |
| DRB1~DQA1~DQB1 | binned | 52.7 | 47.9 | 0.82 (0.43-1.58) | 0.562 |
| DQA1~DQB1 | 01:03:01~06:01:01 | 17.1 | 20.8 | 1.27 (0.50-3.05) | 0.562 |
| DQA1~DQB1 | 03:01:01~03:02:01 | 12.3 | 10.4 | 0.83 (0.23-2.50) | 0.722 |
| DQA1~DQB1 | 03:02:01~03:03:02 | 16.4 | 6.3 | 0.34 (0.06-1.20) | 0.077 |
| DQA1~DQB1 | 01:02:01~06:02:01 | 8.2 | 10.4 | 1.30 (0.43-3.89) | 0.641 |
| DQA1~DQB1 | 03:03:01~04:01:01 | 7.5 | 16.7 | 2.45 (0.92-6.52) | 0.072 |
| DQA1~DQB1 | binned | 38.4 | 35.4 | 0.88 (0.45-1.74) | 0.715 |

IgG-S levels declined after the second dose of BNT162b2 (day 61, 39 days after the second vaccination), showing a significant correlation with age (r=0.325, P=0.012) (Supplementary Figure 2a). DQB1*06:01:01:01 (P = 0.028, OR 0.27, 95%CI 0.05–0.94) was significantly associated with IgG-S declines after two doses of BNT162b2 (Table 4). Haplotype analysis of DQB1*06:01:01:01 revealed significant HLA haplotypes with DQA1, DQA1*01:03:01-DQB1*06:01:01 (P = 0.028; OR 0.27, 95%CI 0.05–0.94). However, the HLA haplotype did not extend to HLA-DRB1 with DRB1*15:02:01-DQB1*06:01:01 (P = 0.392; OR 0.57, 95%CI 0.16–2.05) or DRB1*15:02-DQA1*01:03:01-DQB1*06:01:01 (P = 0.392; OR 0.57, 95%CI 0.16–2.05) (Table 5). Due to the limited sample size, none of the abovementioned HLA alleles remained significant associated with IgG-S levels after multiple correction, but a potential correlation between individual HLA alleles and responsiveness to BNT162b2 vaccination was still suggested.

**Table 4.**
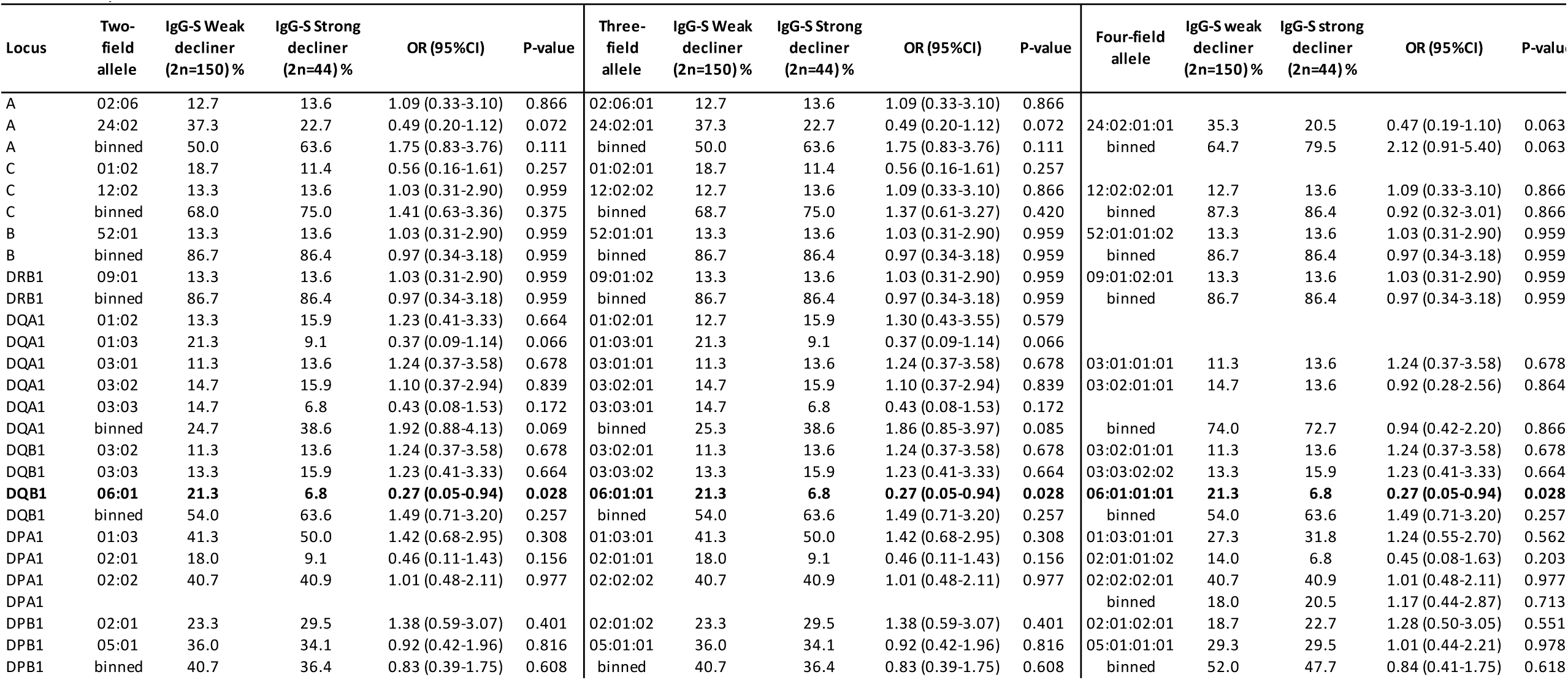
Associational analysis of HLA-A, -C, -B, -DRB1, -DQB1, -DOA1 and -DPB1 alleles in 75 IgG-S weak decliner and 22 IgG-S strong decliner. Abbreviations: OR, odds ratio; 95%CI, 95% confidence interval; binned, rare HLA alleles with expected count < 5 are combined into a common class.

**Table 5.**
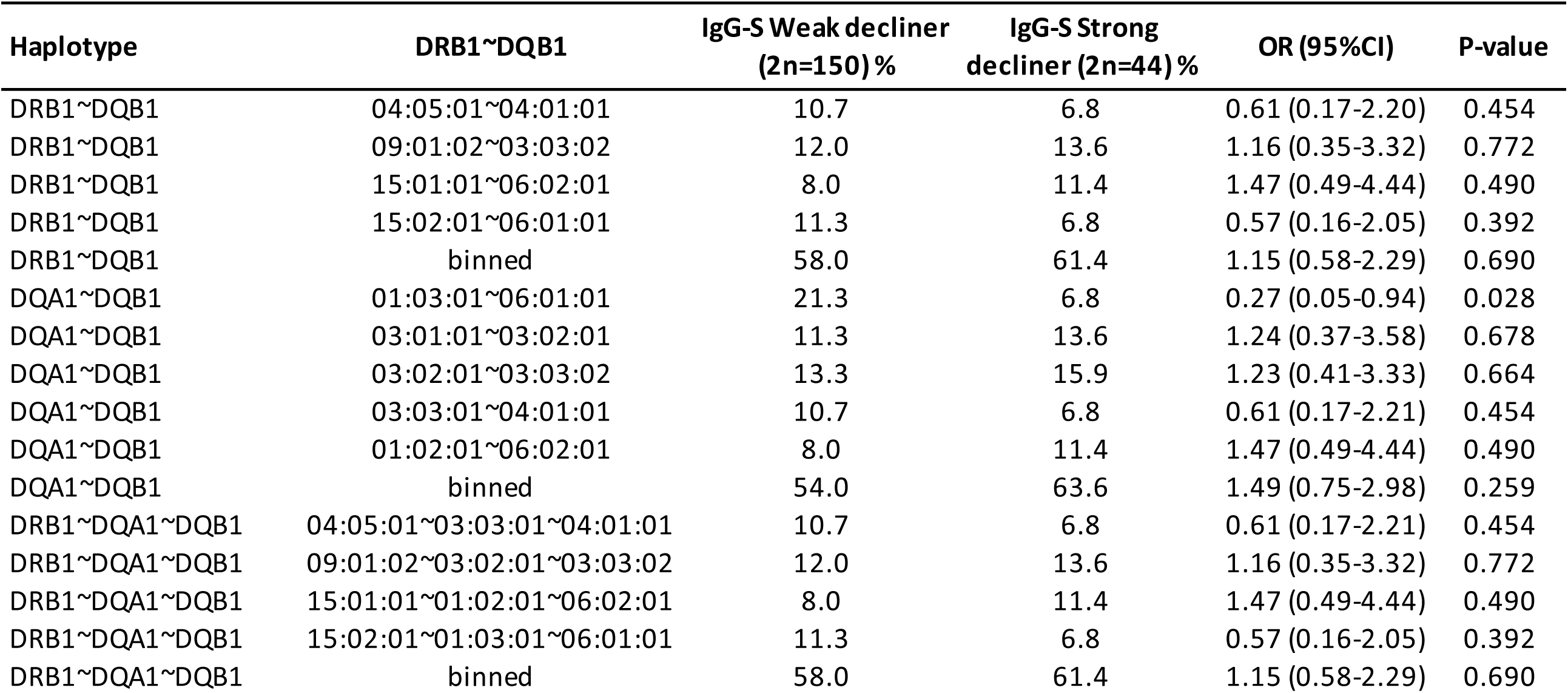
HLA haplotype analysis for HLA-DRB1, -DQA1, and -DQB1 in 75 IgG-S weak decliners and 22 IgG-S strong decliners. Abbreviations: OR, odds ratio; 95%CI, 95% confidence interval; binned, rare HLA alleles with expected count < 5 are combined into a common class.

No HLA alleles were significantly associated with either local symptoms (Supplementary table 1) or fever (Supplementary table 2). However, C*12:02:02 (P = 0.058; OR 0.42, 95%CI 0.15–1.16), B*52:01:01 (P = 0.031; OR 0.38, 95%CI 0.14–1.03), DQA1*03:02:01 (P = 0.028; OR 0.39, 95%CI 0.15–1.00) and DPB1*02:01:02 (P = 0.024; OR 0.45, 95%CI 0.21–0.97) appeared significantly associated with protection against systemic symptoms after two doses of BNT162b2 vaccination (Supplementary table 3).

## Discussion

We have described in this study the kinetic changes in IgG-S profiles and adverse reactions and their associations with HLA profiles among hospital workers from a national medical institution in Japan. BNT162b2 was designed to present the entire spike glycoprotein of SARS-CoV-2 as a target to elicit neutralizing antibodies and so block viral entry through the angiotensin-converting enzyme 2 (ACE2) cellular receptor, particularly the viral sequences recognizing the receptor-binding domain (28). A structural study of SARS-CoV-2 proteins identified the S-protein as a major inducer of protective immunity (30). Assessment of changes in the kinetics of antibodies, adverse reactions after BNT162b2 vaccination, and possible associations between interpersonal differences and variations in HLA genes are thus important.

All participants in the present study displayed seroconversion on day 29 (7 days after the second dose) and displayed an increase of IgG-S against SARS-CoV-2. The youngest age group and female participants produced higher levels of IgG-S after two doses of BNT162b2 vaccinations, consistent with findings from previous studies (13, 31, 32). However, the IgG-S antibody response appears both time- and age-dependent (Figure 2) (11, 13, 16, 33). In this study, the female group produced significantly more IgG-S at the early stage of our longitudinal study, but we also observed a strong IgG-S decline (Δ 20,000 AU/mL) in the female group. DQA1*03:03:01 (P = 0.017; OR 2.80, 95%CI 1.05–7.25) was significantly associated with strong responders for IgG-S, Interestingly, DQA1*03:03:01 is the second most common DQA1 allele in the Japanese population, with a frequency of 16.50% (34).

The long-term sustainability of IgG-S after COVID-19 vaccination is central to the protection afforded against SARS-CoV-2. This study did not identify any HLA alleles associated with strong declines in IgG-S levels. Instead, we identified an association between DQB1*06:01:01:01 (P = 0.028; OR 0.27, 95%CI 0.05–0.94) and protection against IgG-S declines.

The severity of systemic adverse symptoms after two doses of BNT162b2 vaccination was associated with the level of IgG-S production, consistent with previous research (35). We observed that carriers of C*12:02:02, B*52:01:01, DQA1*03:02:01 and DPB1*02:01:02 showed significant protection against systemic symptoms.

Data regarding long-term antibody kinetics in vaccinated subjects remain scarce. In a population of 33 healthy adults having received the Moderna mRNA-1273 vaccine and followed-up for 209 days, the estimated half-life of the antibody response was 52 days (95%CI 46–58 days) using an exponential decay model [13]. In a cohort of 188 unvaccinated COVID-19 patients (mostly not hospitalized: 174/188) who were followed-up for as long as 8 months, the antibody half-life was 83 days (95%CI 62–126 days) [14]. In this study, we estimated that IgG-S levels would drop below 4,160 AU/mL after 101 days, suggesting a need for booster shots to sustain adequate levels of neutralizing antibody.

Research into the associations between HLA alleles and IgG-S antibody levels after vaccination remain limited, but a small Italian cohort (n=56) found no associations between HLA alleles and IgG-S levels after BNT162b2 vaccination. In the present study, we identified several HLA alleles showing potential associations with the kinetics of IgG-S antibody levels. Further studies with larger sample sizes are clearly warranted to determine HLA allele associations with the production and long-term sustainability of IgG-S antibody levels after COVID-19 vaccination.

## Data Availability

All data produced in the present work are contained in the manuscript

## Acknowledgments

We are grateful to the members of the working group for this study (Yoshimi Shigemori, Ayumi Nakayama, Yusuke Oshiro, Natsumi Inamura, Haruka Osawa, Maki Konishi, Azusa Kamikawa, and Yumiko Kito) for their support. We thank Dr. Ikue Ito and Dr. Tetsuya Sato from H.U. Group Research Institute for their technical support for Pacbio Sequel sequencing.

## Figures legends

**Supplementary Figure 1 SARS-CoV-2 spike-specific IgG production on day 29, stratified by age and sex**

a) SARS-CoV-2 spike-specific IgG production on day 29, stratified by 5 age groups: 20–29 years old (y.o), 30–39 y.o., 40–49 y.o. 50–59 y.o, and over 60 y.o

b) SARS-CoV-2 spike-specific IgG production on day 29, stratified by sex

c) SARS-CoV-2 spike-specific IgG production on day 29, stratified by 10 groups including the categories listed in (a) and (b)

Violin plots showing median and 95% confidence interval (CI)

**Supplementary Figure 2 SARS-CoV-2 spike-specific IgG decline on day 61, stratified by age, sex, and BMI of participants**

a) Scatter plot showing the correlation between age and the change in IgG-S between day 29 and day 61. Correlation coefficients and P-values are calculated. Color intensity of the heatmap indicates the accumulation of data points in a specific area.

b) Violin plot showing the correlation between sex and the change in IgG-S between day 29 and day 61. Violin plots showing median and 95% confidence interval (CI)

c) Scatter plot showing the correlation between the BMI of participants and the change in IgG-S between day 29 and day 61. Correlation coefficients and p-values are calculated. Color intensity of the heatmap indicates the accumulation of data points in a specific area

**Supplementary Figure 3 Association between SARS-CoV-2 spike-specific IgG production on day 29 and adverse reactions after two doses of BNT162b2 vaccination**

a) Violin plot showing the distribution of IgG-S production on day 29 stratified by grades of systemic symptoms experienced by participants after two doses of BNT162b2 vaccination

b) Violin plot showing the distribution of IgG-S production on day 29 stratified by fever experienced by participants after two doses of BNT162b2 vaccination. T1: < 37.5°C; T2: 37.5–37.9°C; T3: 38.0–38.4°C; and T4: 38.5–38.9°C.

c) Violin plot showing the distribution of IgG-S production on day 29 stratified by the grade of local symptoms experienced by participants after two doses of BNT162b2 vaccination Violin plots showing median and 95% confidence interval (CI)

