## Supplementary Figure for "Association study of HLA with the kinetics of SARS-CoV-2 spike specific IgG antibody responses to BNT162b2 mRNA vaccine"

**Figures legends**

**Supplementary Figure 1 SARS-CoV-2 spike-specific IgG production on day 29, stratified by age and sex**

a) SARS-CoV-2 spike-specific IgG production on day 29, stratified by 5 age groups: 20–29 years old (y.o), 30–39 y.o., 40–49 y.o. 50–59 y.o, and over 60 y.o

b) SARS-CoV-2 spike-specific IgG production on day 29, stratified by sex

c) SARS-CoV-2 spike-specific IgG production on day 29, stratified by 10 groups including the categories listed in (a) and (b)

Violin plots showing median and 95% confidence interval (CI)

**Supplementary Figure 2 SARS-CoV-2 spike-specific IgG decline on day 61, stratified by age, sex, and BMI of participants**

a) Scatter plot showing the correlation between age and the change in IgG-S between day 29 and day 61. Correlation coefficients and P-values are calculated. Color intensity of the heatmap indicates the accumulation of data points in a specific area.

b) Violin plot showing the correlation between sex and the change in IgG-S between day 29 and day 61. Violin plots showing median and 95% confidence interval (CI)

c) Scatter plot showing the correlation between the BMI of participants and the change in IgG-S between day 29 and day 61. Correlation coefficients and p-values are calculated. Color intensity of the heatmap indicates the accumulation of data points in a specific area

**Supplementary Figure 3 Association between SARS-CoV-2 spike-specific IgG production on day 29 and adverse reactions after two doses of BNT162b2 vaccination**

a) Violin plot showing the distribution of IgG-S production on day 29 stratified by grades of systemic symptoms experienced by participants after two doses of BNT162b2 vaccination

b) Violin plot showing the distribution of IgG-S production on day 29 stratified by fever experienced by participants after two doses of BNT162b2 vaccination. T1: < 37.5°C; T2: 37.5–37.9°C; T3: 38.0–38.4°C; and T4: 38.5–38.9°C.

c) Violin plot showing the distribution of IgG-S production on day 29 stratified by the grade of local symptoms experienced by participants after two doses of BNT162b2 vaccination

Violin plots showing median and 95% confidence interval (CI)

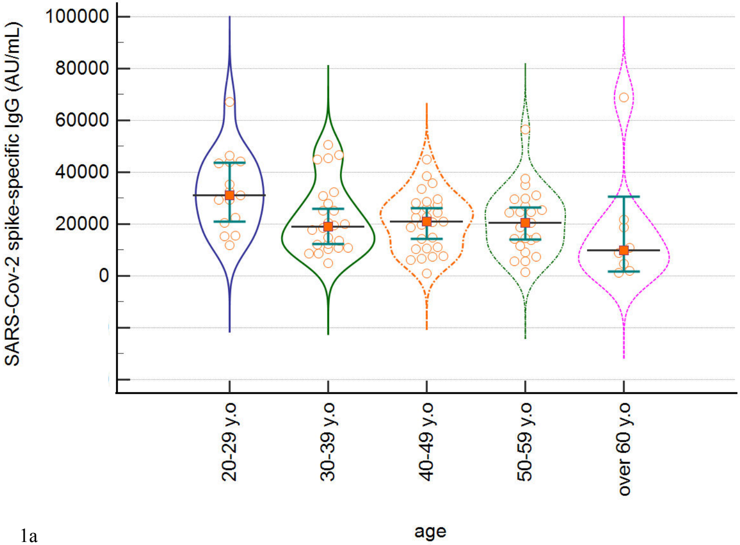
SARS-Cov-2 spike-specific IgG (AU/mL)
100000
80000
60000
40000
20000
0
20-29 y.o
30-39 y.o
40-49 y.o
50-59 y.o
over 60 y.o
age

1a

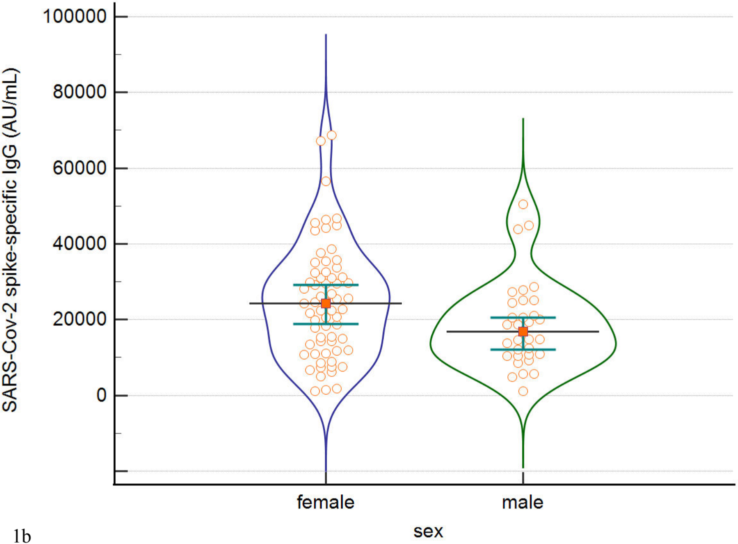
100000
80000
60000
40000
20000
0
SARS-Cov-2 spike-specific IgG (AU/mL)
female
male
sex

1b

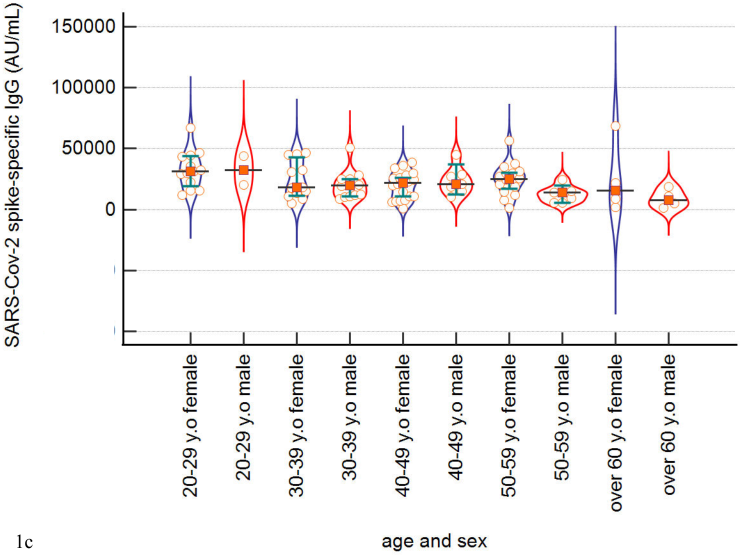
SARS-Cov-2 spike-specific IgG (AU/mL)
150000
100000
50000
0
20-29 y.o female
20-29 y.o male
30-39 y.o female
30-39 y.o male
40-49 y.o female
40-49 y.o male
50-59 y.o female
50-59 y.o male
over 60 y.o female
over 60 y.o male
age and sex

1c

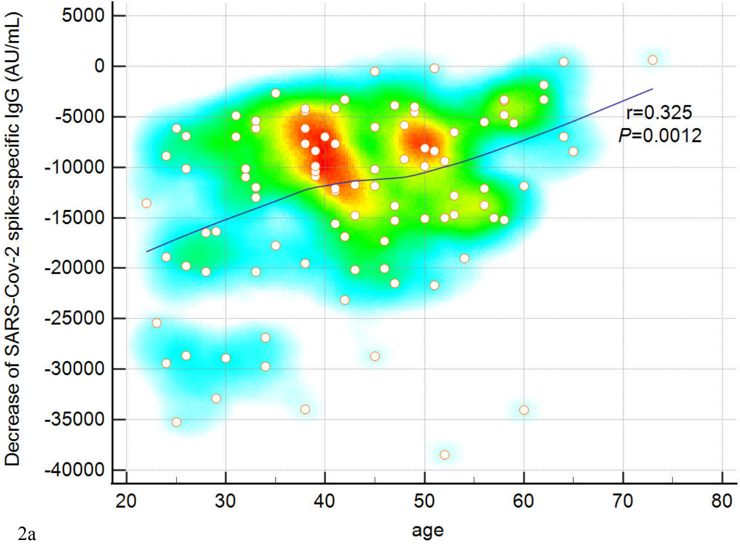

Decrease of SARS-Cov-2 spike-specific IgG (AU/mL)
5000
0
-5000
-10000
-15000
-20000
-25000
-30000
-35000
-40000
20
30
40
50
60
70
80
age
r=0.325
P=0.0012


2a

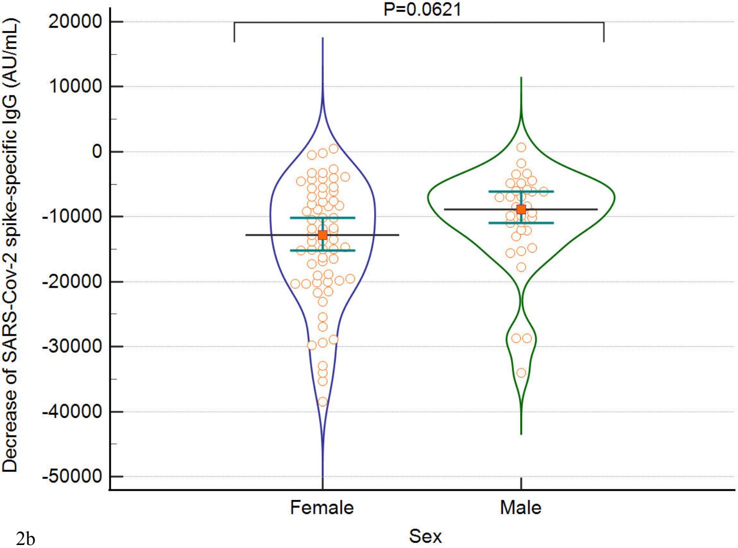
P=0.0621
20000
10000
0
-10000
-20000
-30000
-40000
-50000
Decrease of SARS-Cov-2 spike-specific IgG (AU/mL)
Female
Male
Sex

2b

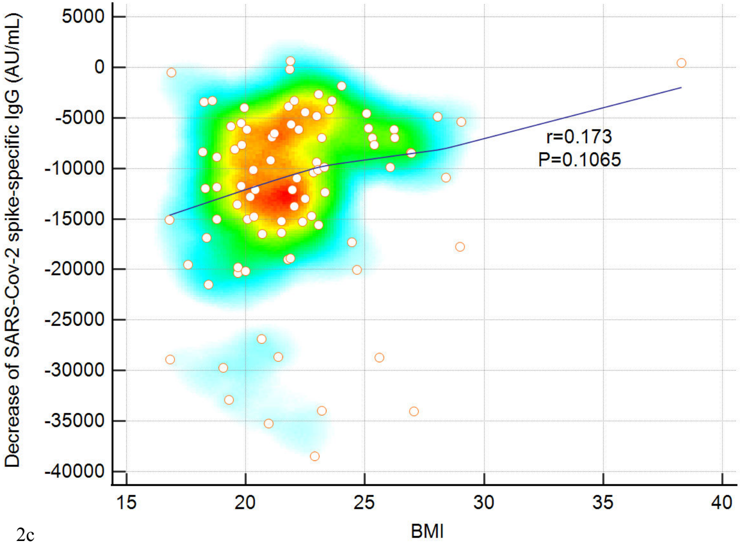

Decrease of SARS-Cov-2 spike-specific IgG (AU/mL)
5000
0
-5000
-10000
-15000
-20000
-25000
-30000
-35000
-40000
15
20
25
30
35
40
BMI
r=0.173
P=0.1065


2c

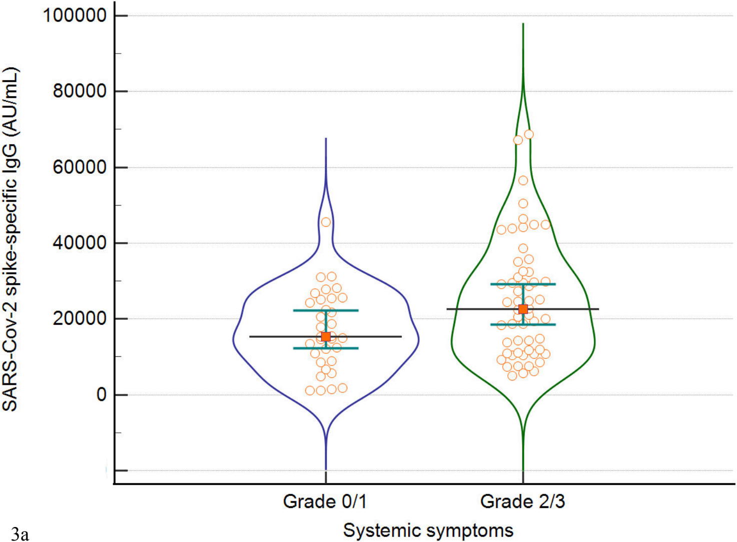

100000
80000
60000
40000
20000
0
SARS-Cov-2 spike-specific IgG (AU/mL)
Grade 0/1
Grade 2/3
Systemic symptoms


3a

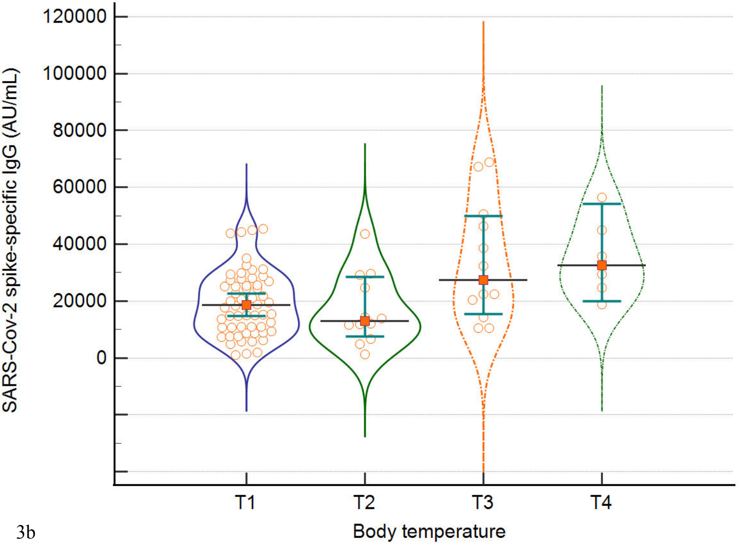
120000
100000
80000
60000
40000
20000
0
SARS-Cov-2 spike-specific IgG (AU/mL)
T1
T2
T3
T4
Body temperature

3b

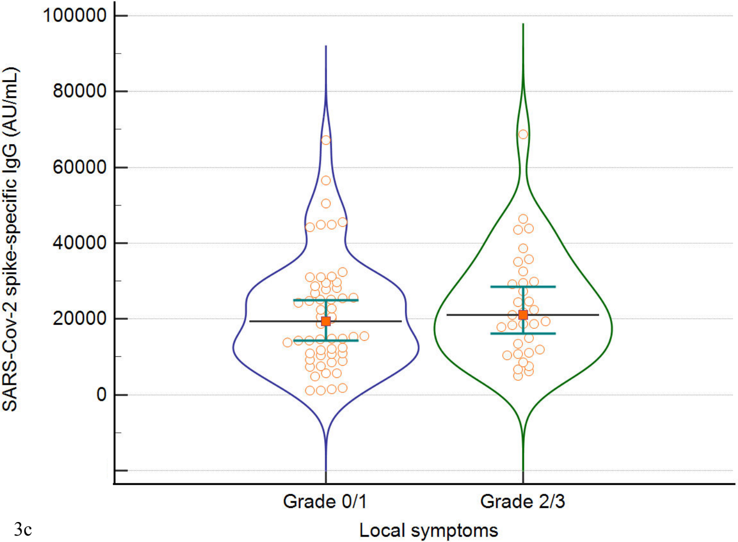
100000
80000
60000
40000
20000
0
SARS-Cov-2 spike-specific IgG (AU/mL)
Grade 0/1
Grade 2/3
Local symptoms
3c
