## Supplementary table for "Association study of HLA with the kinetics of SARS-CoV-2 spike specific IgG antibody responses to BNT162b2 mRNA vaccine"

**Supplementary Table 1 | Associational analysis of HLA-A, -C, -B, -DRB1, -DQB1, -DOA1 and -DPB1 alleles in 55 participants with no local adverse effect versus 33 participants with local adverse effect**

| Locus | Two-field allele | No adverse effect (2n=110) % | Adverse effect (2n=66) % | OR (95%CI) | P-value | Three-field allele | No adverse effect (2n=110) % | Adverse effect (2n=66) % | OR (95%CI) | P-value | Four-field allele | No adverse effect (2n=110) % | Adverse effect (2n=66) % | OR (95%CI) | P-value |
|---|---|---|---|---|---|---|---|---|---|---|---|---|---|---|---|
| A | 02:06 | 15.5 | 10.6 | 0.65 (0.21-1.77) | 0.364 | 02:06:01 | 15.5 | 10.6 | 0.65 (0.21-1.77) | 0.364 | 02:06:01:01 | 11.8 | 6.1 | 0.48 (0.11-1.66) | 0.211 |
| A | 11:01 | 10.9 | 10.6 | 0.97 (0.31-2.85) | 0.950 | 11:01:01 | 10.9 | 10.6 | 0.97 (0.31-2.85) | 0.950 | 11:01:01:01 | 10.9 | 10.6 | 0.97 (0.31-2.85) | 0.950 |
| A | 24:02 | 34.5 | 34.8 | 1.01 (0.50-2.01) | 0.967 | 24:02:01 | 34.5 | 34.8 | 1.01 (0.50-2.01) | 0.967 | 24:02:01:01 | 32.7 | 33.3 | 1.03 (0.51-2.06) | 0.934 |
| A | 31:01 | 7.3 | 10.6 | 1.51 (0.44-5.03) | 0.443 | 31:01:02 | 7.3 | 10.6 | 1.51 (0.44-5.03) | 0.443 | 31:01:02:01 | 6.4 | 10.6 | 1.75 (0.49-6.13) | 0.314 |
| A | binned | 31.8 | 33.3 | 1.07 (0.53-2.15) | 0.835 | binned | 31.8 | 33.3 | 1.07 (0.53-2.15) | 0.835 | binned | 38.2 | 39.4 | 1.05 (0.53-2.06) | 0.873 |
| C | 01:02 | 16.4 | 16.7 | 1.02 (0.40-2.48) | 0.958 | 01:02:01 | 16.4 | 16.7 | 1.02 (0.40-2.48) | 0.958 | 01:02:01:01 | 12.7 | 7.6 | 0.56 (0.15-1.76) | 0.286 |
| C | 03:03 | 9.1 | 15.2 | 1.79 (0.62-5.09) | 0.220 | 03:03:01 | 9.1 | 15.2 | 1.79 (0.62-5.09) | 0.220 | 03:03:01:01 | 9.1 | 15.2 | 1.79 (0.62-5.09) | 0.220 |
| C | 03:04 | 12.7 | 6.1 | 0.44 (0.10-1.50) | 0.158 | 03:04:01 | 12.7 | 6.1 | 0.44 (0.10-1.50) | 0.158 | 03:04:01:02 | 12.7 | 6.1 | 0.44 (0.10-1.50) | 0.158 |
| C | 07:02 | 10.9 | 9.1 | 0.82 (0.24-2.50) | 0.700 | 07:02:01 | 10.9 | 9.1 | 0.82 (0.24-2.50) | 0.700 | | | | | |
| C | 08:01 | 8.2 | 9.1 | 1.12 (0.31-3.73) | 0.834 | 08:01:01 | 8.2 | 9.1 | 1.12 (0.31-3.73) | 0.834 | 08:01:01:01 | 8.2 | 9.1 | 1.12 (0.31-3.73) | 0.834 |
| C | 12:02 | 11.8 | 15.2 | 1.33 (0.49-3.53) | 0.525 | 12:02:02 | 10.9 | 15.2 | 1.46 (0.53-3.95) | 0.410 | 12:02:02:01 | 10.9 | 15.2 | 1.46 (0.53-3.95) | 0.410 |
| C | binned | 30.9 | 28.8 | 0.90 (0.43-1.85) | 0.766 | binned | 31.8 | 28.8 | 0.87 (0.42-1.77) | 0.673 | binned | 46.4 | 47.0 | 1.02 (0.53-1.98) | 0.938 |
| B | 35:01 | 9.1 | 10.6 | 1.19 (0.36-3.67) | 0.742 | 35:01:01 | 9.1 | 10.6 | 1.19 (0.36-3.67) | 0.742 | | | | | |
| B | 40:02 | 11.8 | 6.1 | 0.48 (0.11-1.66) | 0.211 | 40:02:01 | 11.8 | 6.1 | 0.48 (0.11-1.66) | 0.211 | 40:02:01:01 | 10.0 | 4.5 | 0.43 (0.07-1.72) | 0.195 |
| B | 51:01 | 10.0 | 9.1 | 0.90 (0.26-2.82) | 0.843 | 51:01:01 | 10.0 | 9.1 | 0.90 (0.26-2.82) | 0.843 | | | | | |
| B | 52:01 | 11.8 | 15.2 | 1.33 (0.49-3.53) | 0.525 | 52:01:01 | 11.8 | 15.2 | 1.33 (0.49-3.53) | 0.525 | 52:01:01:02 | 11.8 | 15.2 | 1.33 (0.49-3.53) | 0.525 |
| B | binned | 57.3 | 59.1 | 1.08 (0.56-2.10) | 0.813 | binned | 57.3 | 59.1 | 1.08 (0.56-2.1) | 0.813 | binned | 78.2 | 80.3 | 1.14 (0.51-2.65) | 0.738 |
| DRB1 | 04:05 | 10.0 | 10.6 | 1.07 (0.33-3.21) | 0.898 | 04:05:01 | 10.0 | 10.6 | 1.07 (0.33-3.21) | 0.898 | 04:05:01:01 | 10.0 | 10.6 | 1.07 (0.33-3.21) | 0.898 |
| DRB1 | 09:01 | 16.4 | 6.1 | 0.33 (0.08-1.07) | 0.045 | 09:01:02 | 16.4 | 6.1 | 0.33 (0.08-1.07) | 0.045 | 09:01:02:01 | 16.4 | 6.1 | 0.33 (0.08-1.07) | 0.045 |
| DRB1 | 15:01 | 9.1 | 12.1 | 1.38 (0.45-4.12) | 0.521 | 15:01:01 | 9.1 | 12.1 | 1.38 (0.45-4.12) | 0.521 | 15:01:01:01 | 9.1 | 10.6 | 1.19 (0.36-3.67) | 0.742 |
| DRB1 | 15:02 | 10.0 | 10.6 | 1.07 (0.33-3.21) | 0.898 | 15:02:01 | 10.0 | 10.6 | 1.07 (0.33-3.21) | 0.898 | 15:02:01:01 | 10.0 | 10.6 | 1.07 (0.33-3.21) | 0.898 |
| DRB1 | binned | 54.5 | 60.6 | 1.28 (0.66-2.51) | 0.432 | binned | 54.5 | 60.6 | 1.28 (0.66-2.51) | 0.432 | binned | 54.5 | 62.1 | 1.37 (0.70-2.68) | 0.325 |
| DQA1 | 01:02 | 13.6 | 15.2 | 1.13 (0.42-2.90) | 0.780 | 01:02:01 | 13.6 | 13.6 | 1.00 (0.36-2.63) | 1.000 | 01:02:01:01 | 9.1 | 10.6 | 1.19 (0.36-3.67) | 0.742 |
| DQA1 | 01:03 | 18.2 | 18.2 | 1.00 (0.41-2.35) | 1.000 | 01:03:01 | 18.2 | 18.2 | 1.00 (0.41-2.35) | 1.000 | 01:03:01:01 | 10.0 | 13.6 | 1.42 (0.49-4.02) | 0.462 |
| DQA1 | 01:04 | 10.0 | 4.5 | 0.43 (0.07-1.72) | 0.195 | 01:04:01 | 10.0 | 4.5 | 0.43 (0.07-1.72) | 0.195 | 01:04:01:01 | 10.0 | 4.5 | 0.43 (0.07-1.72) | 0.195 |
| DQA1 | 03:01 | 11.8 | 13.6 | 1.18 (0.42-3.19) | 0.724 | 03:01:01 | 11.8 | 13.6 | 1.18 (0.42-3.19) | 0.724 | 03:01:01:01 | 11.8 | 13.6 | 1.18 (0.42-3.19) | 0.724 |
| DQA1 | 03:02 | 18.2 | 7.6 | 0.37 (0.10-1.09) | 0.051 | 03:02:01 | 18.2 | 7.6 | 0.37 (0.10-1.09) | 0.051 | 03:02:01:01 | 17.3 | 7.6 | 0.39 (0.11-1.17) | 0.070 |
| DQA1 | 03:03 | 13.6 | 12.1 | 0.87 (0.30-2.36) | 0.773 | 03:03:01 | 13.6 | 12.1 | 0.87 (0.30-2.36) | 0.773 | 03:03:01:03 | 9.1 | 10.6 | 1.19 (0.36-3.67) | 0.742 |
| DQA1 | binned | 14.5 | 28.8 | 2.38 (1.05-5.41) | 0.022 | binned | 14.5 | 30.3 | 2.55 (1.13-5.79) | 0.012 | binned | 32.7 | 39.4 | 1.34 (0.67-2.64) | 0.370 |
| DQB1 | 03:01 | 9.1 | 15.2 | 1.79 (0.62-5.09) | 0.220 | 03:01:01 | 9.1 | 15.2 | 1.79 (0.62-5.09) | 0.220 | | | | | |
| DQB1 | 03:02 | 11.8 | 13.6 | 1.18 (0.42-3.19) | 0.724 | 03:02:01 | 11.8 | 13.6 | 1.18 (0.42-3.19) | 0.724 | 03:02:01:01 | 11.8 | 13.6 | 1.18 (0.42-3.19) | 0.724 |
| DQB1 | 03:03 | 17.3 | 7.6 | 0.39 (0.11-1.17) | 0.070 | 03:03:02 | 17.3 | 7.6 | 0.39 (0.11-1.17) | 0.070 | 03:03:02:02 | 17.3 | 7.6 | 0.39 (0.11-1.17) | 0.070 |
| DQB1 | 04:01 | 9.1 | 10.6 | 1.19 (0.36-3.67) | 0.742 | 04:01:01 | 9.1 | 10.6 | 1.19 (0.36-3.67) | 0.742 | 04:01:01:01 | 9.1 | 10.6 | 1.19 (0.36-3.67) | 0.742 |
| DQB1 | 06:01 | 18.2 | 16.7 | 0.90 (0.36-2.15) | 0.798 | 06:01:01 | 18.2 | 16.7 | 0.90 (0.36-2.15) | 0.798 | 06:01:01:01 | 18.2 | 16.7 | 0.90 (0.36-2.15) | 0.798 |
| DQB1 | 06:02 | 9.1 | 10.6 | 1.19 (0.36-3.67) | 0.742 | 06:02:01 | 9.1 | 10.6 | 1.19 (0.36-3.67) | 0.742 | 06:02:01:01 | 9.1 | 10.6 | 1.19 (0.36-3.67) | 0.742 |
| DQB1 | binned | 25.5 | 25.8 | 1.02 (0.47-2.15) | 0.964 | binned | 25.5 | 25.8 | 1.02 (0.47-2.15) | 0.964 | binned | 34.5 | 40.9 | 1.31 (0.66-2.57) | 0.397 |
| DPA1 | 01:03 | 43.6 | 40.9 | 0.89 (0.46-1.73) | 0.723 | 01:03:01 | 43.6 | 40.9 | 0.89 (0.46-1.73) | 0.723 | 01:03:01:01 | 30.9 | 22.7 | 0.66 (0.30-1.39) | 0.241 |
| DPA1 | | | | | | | | | | | 01:03:01:05 | 9.1 | 13.6 | 1.58 (0.53-4.60) | 0.347 |
| DPA1 | 02:01 | 14.5 | 21.2 | 1.58 (0.66-3.76) | 0.255 | 02:01:01 | 14.5 | 21.2 | 1.58 (0.66-3.76) | 0.255 | 02:01:01:02 | 9.1 | 19.7 | 2.45 (0.92-6.67) | 0.043 |
| DPA1 | 02:02 | 41.8 | 37.9 | 0.85 (0.43-1.66) | 0.606 | 02:02:02 | 41.8 | 37.9 | 0.85 (0.43-1.66) | 0.606 | 02:02:02:01 | 41.8 | 37.9 | 0.85 (0.43-1.66) | 0.606 |
| DPA1 | | | | | | | | | | | binned | 9.1 | 6.1 | 0.65 (0.14-2.36) | 0.472 |
| DPB1 | 02:01 | 26.4 | 18.2 | 0.62 (0.27-1.39) | 0.214 | 02:01:02 | 26.4 | 18.2 | 0.62 (0.27-1.39) | 0.214 | 02:01:02:01 | 20.9 | 15.2 | 0.68 (0.27-1.61) | 0.343 |
| DPB1 | 04:02 | 9.1 | 13.6 | 1.58 (0.53-4.60) | 0.347 | 04:02:01 | 9.1 | 13.6 | 1.58 (0.53-4.60) | 0.347 | 04:02:01:02 | 9.1 | 12.1 | 1.38 (0.45-4.12) | 0.521 |
| DPB1 | 05:01 | 36.4 | 37.9 | 1.07 (0.54-2.10) | 0.840 | 05:01:01 | 36.4 | 37.9 | 1.07 (0.54-2.10) | 0.840 | 05:01:01:01 | 31.8 | 28.8 | 0.87 (0.42-1.77) | 0.673 |
| DPB1 | 09:01 | 7.3 | 15.2 | 2.28 (0.76-7.02) | 0.095 | 09:01:01 | 7.3 | 15.2 | 2.28 (0.76-7.02) | 0.095 | 09:01:01 | 7.3 | 15.2 | 2.28 (0.76-7.02) | 0.095 |
| DPB1 | binned | 20.9 | 15.2 | 0.68 (0.27-1.61) | 0.343 | binned | 20.9 | 15.2 | 0.68 (0.27-1.61) | 0.343 | binned | 30.9 | 28.8 | 0.90 (0.43-1.85) | 0.766 |

Supplementary Table 2 | Associational analysis of HLA-A, -C, -B, -DRB1, -DQB1, -DOA1 and -DPB1 alleles in 70 participants with fever versus 18 participants with fever more than 38°C

| Locus | Two-field allele | No fever (2n=140) % | Fever (>38°C) (2n=36) % | OR (95%CI) | p-value | Three-field allele | No fever (2n=140) % | Fever (>38°C) (2n=36) % | OR (95%CI) | p-value | Four-field allele | No fever (2n=140) % | Fever (>38°C) (2n=36) % | OR (95%CI) | p-value |
|---|---|---|---|---|---|---|---|---|---|---|---|---|---|---|---|
| A | 02:06 | 13.6 | 16.7 | 1.27 (0.38-3.69) | 0.635 | 02:06:01 | 13.6 | 16.7 | 1.27 (0.38-3.69) | 0.635 | | | | | |
| A | 24:02 | 36.4 | 27.8 | 0.67 (0.27-1.58) | 0.331 | 24:02:01 | 36.4 | 27.8 | 0.67 (0.27-1.58) | 0.331 | 24:02:01:01 | 33.6 | 27.8 | 0.76 (0.30-1.80) | 0.508 |
| A | binned | 50.0 | 55.6 | 1.25 (0.56-2.81) | 0.552 | binned | 50.0 | 55.6 | 1.25 (0.56-2.81) | 0.552 | binned | 66.4 | 72.2 | 1.31 (0.55-3.31) | 0.508 |
| C | 01:02 | 17.1 | 13.9 | 0.78 (0.22-2.32) | 0.639 | 01:02:01 | 17.1 | 13.9 | 0.78 (0.22-2.32) | 0.639 | | | | | |
| C | binned | 82.9 | 86.1 | 1.28 (0.43-4.65) | 0.639 | binned | 82.9 | 86.1 | 1.28 (0.43-4.65) | 0.639 | binned | 100.0 | 100.0 | NA | NA |
| B | binned | 100.0 | 100.0 | NA | NA | binned | 100.0 | 100.0 | NA | NA | binned | 100.0 | 100.0 | NA | NA |
| DRB1 | binned | 100.0 | 100.0 | NA | NA | binned | 100.0 | 100.0 | NA | NA | binned | 100.0 | 100.0 | NA | NA |
| DQA1 | 01:02 | 13.6 | 16.7 | 1.27 (0.38-3.69) | 0.635 | | | | | | | | | | |
| DQA1 | 01:03 | 20.7 | 11.1 | 0.48 (0.11-1.52) | 0.188 | 01:03:01 | 20.7 | 11.1 | 0.48 (0.11-1.52) | 0.188 | | | | | |
| DQA1 | 03:02 | 15.0 | 11.1 | 0.71 (0.17-2.32) | 0.551 | 03:02:01 | 15.0 | 11.1 | 0.71 (0.17-2.32) | 0.551 | | | | | |
| DQA1 | binned | 50.7 | 61.1 | 1.53 (0.68-3.50) | 0.265 | binned | 64.3 | 77.8 | 1.94 (0.78-5.3) | 0.125 | binned | 100.0 | 100.0 | NA | NA |
| DQB1 | 06:01 | 20.0 | 11.1 | 0.50 (0.12-1.59) | 0.217 | 06:01:01 | 20.0 | 11.1 | 0.50 (0.12-1.59) | 0.217 | 06:01:01:01 | 20.0 | 11.1 | 0.50 (0.12-1.59) | 0.217 |
| DQB1 | binned | 80.0 | 88.9 | 2.00 (0.63-8.39) | 0.217 | binned | 80.0 | 88.9 | 2.00 (0.63-8.39) | 0.217 | binned | 80.0 | 88.9 | 2.00 (0.63-8.39) | 0.217 |
| DPA1 | 01:03 | 42.9 | 38.9 | 0.85 (0.37-1.90) | 0.667 | 01:03:01 | 42.9 | 38.9 | 0.85 (0.37-1.90) | 0.667 | 01:03:01:01 | 28.6 | 25.0 | 0.83 (0.32-2.03) | 0.670 |
| DPA1 | 02:01 | 17.1 | 16.7 | 0.97 (0.30-2.72) | 0.946 | 02:01:01 | 17.1 | 16.7 | 0.97 (0.30-2.72) | 0.946 | | | | | |
| DPA1 | 02:02 | 40.0 | 44.4 | 1.20 (0.53-2.67) | 0.629 | 02:02:02 | 40.0 | 44.4 | 1.20 (0.53-2.67) | 0.629 | 02:02:02:01 | 40.0 | 44.4 | 1.20 (0.53-2.67) | 0.629 |
| DPA1 | | | | | | | | | | | binned | 31.4 | 30.6 | 0.96 (0.39-2.24) | 0.920 |
| DPB1 | 02:01 | 24.3 | 19.4 | 0.75 (0.26-1.97) | 0.540 | 02:01:02 | 24.3 | 19.4 | 0.75 (0.26-1.97) | 0.540 | 02:01:02:01 | 18.6 | 19.4 | 1.06 (0.35-2.83) | 0.905 |
| DPB1 | 05:01 | 36.4 | 38.9 | 1.11 (0.48-2.50) | 0.785 | 05:01:01 | 36.4 | 38.9 | 1.11 (0.48-2.50) | 0.785 | 05:01:01:01 | 31.4 | 27.8 | 0.84 (0.33-1.99) | 0.672 |
| DPB1 | binned | 39.3 | 41.7 | 1.10 (0.48-2.47) | 0.795 | binned | 39.3 | 41.7 | 1.10 (0.48-2.47) | 0.795 | binned | 50.0 | 52.8 | 1.12 (0.50-2.50) | 0.766 |

**Supplementary Table 3 | Associational analysis of HLA-A, -C, -B, -DRB1, -DQB1, -DOA1 and -DPB1 alleles in 55 participants with no systemic adverse effect versus 33 participants with systemic adverse effect**

| Locus | Two-field allele | No adverse effect (n=64) % | With adverse effect (n=112) % | OR (95%CI) | p-value | Three-field allele | No adverse effect (n=64) % | With adverse effect (n=112) % | OR (95%CI) | p-value | Four-field allele | No adverse effect (n=64) % | With adverse effect (n=112) % | OR (95%CI) | p-value |
|---|---|---|---|---|---|---|---|---|---|---|---|---|---|---|---|
| A | 02:06 | 15.6 | 12.5 | 0.77 (0.30-2.09) | 0.561 | 02:06:01 | 15.6 | 12.5 | 0.77 (0.30-2.09) | 0.561 | 02:06:01:01 | 12.5 | 8.0 | 0.61 (0.20-1.94) | 0.335 |
| A | 11:01 | 10.9 | 10.7 | 0.98 (0.33-3.11) | 0.963 | 11:01:01 | 10.9 | 10.7 | 0.98 (0.33-3.11) | 0.963 | 11:01:01:01 | 10.9 | 10.7 | 0.98 (0.33-3.11) | 0.963 |
| A | 24:02 | 35.9 | 33.9 | 0.92 (0.46-1.84) | 0.788 | 24:02:01 | 35.9 | 33.9 | 0.92 (0.46-1.84) | 0.788 | 24:02:01:01 | 34.4 | 32.1 | 0.90 (0.45-1.84) | 0.762 |
| A | 31:01 | 10.9 | 7.1 | 0.63 (0.19-2.15) | 0.386 | 31:01:02 | 10.9 | 7.1 | 0.63 (0.19-2.15) | 0.386 | 31:01:02:01 | 9.4 | 7.1 | 0.74 (0.21-2.74) | 0.599 |
| A | binned | 26.6 | 35.7 | 1.54 (0.75-3.23) | 0.212 | binned | 26.6 | 35.7 | 1.54 (0.75-3.23) | 0.212 | binned | 32.8 | 42.0 | 1.48 (0.74-2.98) | 0.230 |
| C | 01:02 | 10.9 | 19.6 | 1.99 (0.76-5.86) | 0.134 | 01:02:01 | 10.9 | 19.6 | 1.99 (0.76-5.86) | 0.134 | 01:02:01:01 | 6.3 | 13.4 | 2.32 (0.69-10.01) | 0.142 |
| C | 03:03 | 18.8 | 7.1 | 0.33 (0.11-0.96) | 0.020 | 03:03:01 | 18.8 | 7.1 | 0.33 (0.11-0.96) | 0.020 | 03:03:01:01 | 18.8 | 7.1 | 0.33 (0.11-0.96) | 0.020 |
| C | 03:04 | 12.5 | 8.9 | 0.69 (0.23-2.13) | 0.452 | 03:04:01 | 12.5 | 8.9 | 0.69 (0.23-2.13) | 0.452 | 03:04:01:02 | 12.5 | 8.9 | 0.69 (0.23-2.13) | 0.452 |
| C | 07:02 | 12.5 | 8.9 | 0.69 (0.23-2.13) | 0.452 | 07:02:01 | 12.5 | 8.9 | 0.69 (0.23-2.13) | 0.452 | | | | | |
| C | 08:01 | 6.3 | 9.8 | 1.63 (0.46-7.33) | 0.414 | 08:01:01 | 6.3 | 9.8 | 1.63 (0.46-7.33) | 0.414 | 08:01:01:01 | 6.3 | 9.8 | 1.63 (0.46-7.33) | 0.414 |
| **C** | **12:02** | **20.3** | **8.9** | **0.38 (0.14-1.03)** | **0.031** | **12:02:02** | **18.8** | **8.9** | **0.42 (0.15-1.16)** | **0.058** | **12:02:02:01** | **18.8** | **8.9** | **0.42 (0.15-1.16)** | **0.058** |
| C | binned | 18.8 | 36.6 | 2.50 (1.15-5.73) | 0.013 | binned | 20.3 | 36.6 | 2.27 (1.05-5.08) | 0.024 | binned | 37.5 | 51.8 | 1.79 (0.91-3.53) | 0.068 |
| B | 35:01 | 12.5 | 8.0 | 0.61 (0.20-1.94) | 0.335 | 35:01:01 | 12.5 | 8.0 | 0.61 (0.20-1.94) | 0.335 | | | | | |
| B | 40:02 | 14.1 | 7.1 | 0.47 (0.15-1.47) | 0.135 | 40:02:01 | 14.1 | 7.1 | 0.47 (0.15-1.47) | 0.135 | 40:02:01:01 | 10.9 | 6.3 | 0.54 (0.15-1.92) | 0.269 |
| B | 51:01 | 6.3 | 11.6 | 1.97 (0.57-8.65) | 0.247 | 51:01:01 | 6.3 | 11.6 | 1.97 (0.57-8.65) | 0.247 | | | | | |
| **B** | **52:01** | **20.3** | **8.9** | **0.38 (0.14-1.03)** | **0.031** | **52:01:01** | **20.3** | **8.9** | **0.38 (0.14-1.03)** | **0.031** | **52:01:01:02** | **20.3** | **8.9** | **0.38 (0.14-1.03)** | **0.031** |
| B | binned | 46.9 | 64.3 | 2.04 (1.04-4.00) | 0.024 | binned | 46.9 | 64.3 | 2.04 (1.04-4.00) | 0.024 | binned | 68.8 | 84.8 | 2.54 (1.13-5.69) | 0.012 |
| DRB1 | 04:05 | 7.8 | 11.6 | 1.55 (0.49-5.82) | 0.424 | 04:05:01 | 7.8 | 11.6 | 1.55 (0.49-5.82) | 0.424 | 04:05:01:01 | 7.8 | 11.6 | 1.55 (0.49-5.82) | 0.424 |
| DRB1 | 09:01 | 18.8 | 8.9 | 0.42 (0.15-1.16) | 0.058 | 09:01:02 | 18.8 | 8.9 | 0.42 (0.15-1.16) | 0.058 | 09:01:02:01 | 18.8 | 8.9 | 0.42 (0.15-1.16) | 0.058 |
| DRB1 | 15:01 | 12.5 | 8.9 | 0.69 (0.23-2.13) | 0.452 | 15:01:01 | 12.5 | 8.9 | 0.69 (0.23-2.13) | 0.452 | 15:01:01:01 | 10.9 | 8.9 | 0.80 (0.26-2.62) | 0.664 |
| DRB1 | 15:02 | 15.6 | 7.1 | 0.42 (0.13-1.25) | 0.074 | 15:02:01 | 15.6 | 7.1 | 0.42 (0.13-1.25) | 0.074 | 15:02:01:01 | 15.6 | 7.1 | 0.42 (0.13-1.25) | 0.074 |
| DRB1 | binned | 45.3 | 63.4 | 2.09 (1.07-4.09) | 0.020 | binned | 45.3 | 63.4 | 2.09 (1.07-4.09) | 0.020 | binned | 46.9 | 63.4 | 1.96 (1.00-3.84) | 0.033 |
| DQA1 | 01:02 | 15.6 | 13.4 | 0.84 (0.33-2.23) | 0.683 | 01:02:01 | 14.1 | 13.4 | 0.95 (0.36-2.62) | 0.901 | 01:02:01:01 | 12.5 | 8.0 | 0.61 (0.20-1.94) | 0.335 |
| DQA1 | 01:03 | 21.9 | 16.1 | 0.68 (0.29-1.62) | 0.337 | 01:03:01 | 21.9 | 16.1 | 0.68 (0.29-1.62) | 0.337 | 01:03:01:01 | 15.6 | 8.9 | 0.53 (0.19-1.52) | 0.178 |
| DQA1 | 01:04 | 6.3 | 8.9 | 1.47 (0.4-6.69) | 0.528 | 01:04:01 | 6.3 | 8.9 | 1.47 (0.40-6.69) | 0.528 | 01:04:01:01 | 6.3 | 8.9 | 1.47 (0.40-6.69) | 0.528 |
| DQA1 | 03:01 | 10.9 | 13.4 | 1.26 (0.45-3.87) | 0.636 | 03:01:01 | 10.9 | 13.4 | 1.26 (0.45-3.87) | 0.636 | 03:01:01:01 | 10.9 | 13.4 | 1.26 (0.45-3.87) | 0.636 |
| **DQA1** | **03:02** | **21.9** | **9.8** | **0.39 (0.15-1.00)** | **0.028** | **03:02:01** | **21.9** | **9.8** | **0.39 (0.15-1.00)** | **0.028** | **03:02:01:01** | **21.9** | **8.9** | **0.35 (0.13-0.92)** | **0.016** |
| DQA1 | 03:03 | 7.8 | 16.1 | 2.26 (0.75-8.17) | 0.118 | 03:03:01 | 7.8 | 16.1 | 2.26 (0.75-8.17) | 0.118 | 03:03:01:03 | 6.3 | 11.6 | 1.97 (0.57-8.65) | 0.247 |
| DQA1 | binned | 15.6 | 22.3 | 1.55 (0.66-3.91) | 0.284 | binned | 17.2 | 22.3 | 1.38 (0.60-3.38) | 0.417 | binned | 26.6 | 40.2 | 1.86 (0.91-3.89) | 0.069 |
| DQB1 | 03:01 | 7.8 | 13.4 | 1.82 (0.59-6.73) | 0.262 | 03:01:01 | 7.8 | 13.4 | 1.82 (0.59-6.73) | 0.262 | | | | | |
| DQB1 | 03:02 | 10.9 | 13.4 | 1.26 (0.45-3.87) | 0.636 | 03:02:01 | 10.9 | 13.4 | 1.26 (0.45-3.87) | 0.636 | 03:02:01:01 | 10.9 | 13.4 | 1.26 (0.45-3.87) | 0.636 |
| DQB1 | 03:03 | 20.3 | 9.8 | 0.43 (0.16-1.12) | 0.051 | 03:03:02 | 20.3 | 9.8 | 0.43 (0.16-1.12) | 0.051 | 03:03:02:02 | 20.3 | 9.8 | 0.43 (0.16-1.12) | 0.051 |
| DQB1 | 04:01 | 6.3 | 11.6 | 1.97 (0.57-8.65) | 0.247 | 04:01:01 | 6.3 | 11.6 | 1.97 (0.57-8.65) | 0.247 | 04:01:01:01 | 6.3 | 11.6 | 1.97 (0.57-8.65) | 0.247 |
| DQB1 | 06:01 | 21.9 | 15.2 | 0.64 (0.27-1.53) | 0.262 | 06:01:01 | 21.9 | 15.2 | 0.64 (0.27-1.53) | 0.262 | 06:01:01:01 | 21.9 | 15.2 | 0.64 (0.27-1.53) | 0.262 |
| DQB1 | 06:02 | 12.5 | 8.0 | 0.61 (0.20-1.94) | 0.335 | 06:02:01 | 12.5 | 8.0 | 0.61 (0.20-1.94) | 0.335 | 06:02:01:01 | 12.5 | 8.0 | 0.61 (0.20-1.94) | 0.335 |
| DQB1 | binned | 20.3 | 28.6 | 1.57 (0.72-3.57) | 0.227 | binned | 20.3 | 28.6 | 1.57 (0.72-3.57) | 0.227 | binned | 28.1 | 42.0 | 1.85 (0.91-3.82) | 0.067 |
| DPA1 | 01:03 | 40.6 | 43.8 | 1.14 (0.58-2.23) | 0.687 | 01:03:01 | 40.6 | 43.8 | 1.14 (0.58-2.23) | 0.687 | 01:03:01:01 | 32.8 | 25.0 | 0.68 (0.33-1.43) | 0.266 |
| DPA1 | | | | | | | | | | | 01:03:01:05 | 4.7 | 14.3 | 3.39 (0.91-18.79) | 0.048 |
| DPA1 | 02:01 | 21.9 | 14.3 | 0.60 (0.25-1.44) | 0.198 | 02:01:01 | 21.9 | 14.3 | 0.60 (0.25-1.44) | 0.198 | 02:01:01:02 | 14.1 | 12.5 | 0.87 (0.33-2.45) | 0.767 |
| DPA1 | 02:02 | 37.5 | 42.0 | 1.21 (0.61-2.39) | 0.561 | 02:02:02 | 37.5 | 42.0 | 1.21 (0.61-2.39) | 0.561 | 02:02:02:01 | 37.5 | 42.0 | 1.21 (0.61-2.39) | 0.561 |
| DPA1 | | | | | | | | | | | binned | 10.9 | 6.3 | 0.54 (0.15-1.92) | 0.269 |
| **DPB1** | **02:01** | **32.8** | **17.9** | **0.45 (0.21-0.97)** | **0.024** | **02:01:02** | **32.8** | **17.9** | **0.45 (0.21-0.97)** | **0.024** | **02:01:02:01** | **25.0** | **15.2** | **0.54 (0.23-1.25)** | **0.108** |
| DPB1 | 04:02 | 4.7 | 14.3 | 3.39 (0.91-18.79) | 0.048 | 04:02:01 | 4.7 | 14.3 | 3.39 (0.91-18.79) | 0.048 | 04:02:01:02 | 4.7 | 13.4 | 3.14 (0.84-17.54) | 0.067 |
| DPB1 | 05:01 | 32.8 | 39.3 | 1.32 (0.66-2.68) | 0.392 | 05:01:01 | 32.8 | 39.3 | 1.32 (0.66-2.68) | 0.392 | 05:01:01:01 | 29.7 | 31.3 | 1.08 (0.53-2.24) | 0.829 |
| DPB1 | 09:01 | 9.4 | 10.7 | 1.16 (0.38-3.97) | 0.778 | 09:01:01 | 9.4 | 10.7 | 1.16 (0.38-3.97) | 0.778 | 09:01:01 | 9.4 | 10.7 | 1.16 (0.38-3.97) | 0.778 |
| DPB1 | binned | 20.3 | 17.9 | 0.85 (0.37-2.03) | 0.688 | binned | 20.3 | 17.9 | 0.85 (0.37-2.03) | 0.688 | binned | 31.3 | 29.5 | 0.92 (0.45-1.91) | 0.804 |
